# Clinical symptoms, cognitive performance and cortical activity following mild traumatic brain injury (mTBI)

**DOI:** 10.1101/2022.06.05.22275980

**Authors:** Hannah L. Coyle, Neil W. Bailey, Jennie Ponsford, Kate E. Hoy

## Abstract

**Objective:** To investigate clinical symptoms, cognitive performance and cortical activity following mild traumatic brain injury (mTBI).

**Methods:** Thirty individuals in the sub-acute phase post mTBI and 28 healthy controls with no history of head injury were compared on clinical, cognitive and cortical activity measures. Measures of cortical activity included; resting state EEG, task related EEG and combined transcranial magnetic stimulation with electroencephalography (TMS-EEG). Primary analyses investigated clinical, cognitive and cortical activity differences between groups. Exploratory analyses investigated the relationships between these measures.

**Results:** At 4 weeks’ post injury, mTBI participants exhibited significantly greater post concussive and clinical symptoms compared to controls; as well as reduced cognitive performance on verbal learning and working memory measures. mTBI participants demonstrated alterations in cortical activity while at rest and in response to stimulation with TMS.

**Conclusions:** The mTBI group demonstrated neurophysiological markers of altered excitatory and inhibitory processes which impact neural function. Further research is needed to explore the relationship between these pathophysiologies and clinical/cognitive symptoms in mTBI.

## 1. Introduction

Traumatic brain injury (TBI) has been estimated to affect 69 million people worldwide every year, with mild traumatic brain injuries (mTBI) accounting for around 80% of all TBIs (Dewan et al., 2018). There is significant heterogeneity in symptoms post mTBI, with some individuals reporting persistent symptoms and others rapidly returning to pre-injury functioning. The factors that contribute to this variability are currently poorly understood. A more nuanced characterisation of how symptoms and pathophysiology interact may aid in our understanding of heterogeneous presentations. Further developing our understanding of the relationships between symptoms and pathophysiology may improve clinical decision-making precision around diagnosis and prognosis, as well as the development of targeted therapeutic interventions.

The relationship between mechanisms of injury and symptoms in mTBI is well established (Karr et al. 2014; Rohling et al. 2011; Xiao et al. 2015). In brief, due to the viscoelastic nature of the brain, bidirectional forces during injury result in shearing of cell membranes and cytoskeletal elements (Giza and Hovda 2014). Excitatory and inhibitory neural circuit imbalances post mTBI, commonly referred to as the “neurometabolic cascade” (Giza and Hovda 2014), are associated with neuronal dysfunction, disrupted connectivity and impaired cortical information processing (Zhou and Yu 2018). Indeed, there has been extensive neuroimaging research showing that clinical and cognitive symptoms correlate with structural and functional connectivity changes following mTBI (e.g. FitzGerald and Crosson 2011; Kinnunen et al. 2011; Mayer et al. 2011; Palacios et al. 2017; Salmond et al. 2006; Yuh et al. 2014).

Frontoparietal regions and subcortical structures with critical frontal projections have been shown to be particularly susceptible to damage following TBI (Eierud et al. 2014; Lipton et al. 2009). Frontal brain regions are implicated in networks relevant to cognition and emotion regulation, offering an explanatory mechanism for the cognitive symptoms that are common after TBI, and highlighting these brain regions as areas of interest. mTBI, however, is not considered a focal injury, as disruptions to neuronal function impact both local and global brain network connectivity (Sharp et al. 2014). The pathophysiological complexity of mTBI highlights the importance of using complementary imaging modalities to understand the neural changes that occur throughout the brain and their relationships to symptoms.

Electroencephalography (EEG) is a non-invasive imaging method that can provide information on brain dynamics with millisecond precision, allowing brain activity to be measured at rest or during cognitive processing. Previous research has detected a range of EEG abnormalities following mTBI, both in the presence and absence of clinical and cognitive symptoms, highlighting the sensitivity of EEG as a measure of disrupted brain function (Broglio et al. 2011; Dockree and Robertson 2011; Haneef et al. 2013; Rapp et al. 2015). Transcranial magnetic stimulation (TMS) has also been used to investigate cortical activity changes post mTBI, however most of this work has been in the motor cortex (Major et al. 2015). While these motor cortex findings have varied considerably across studies, likely due to methodological variability, changes have been detected in a range of TMS components that reflect cortical reactivity (Chistyakov et al. 2001; De Beaumont et al. 2007; De Beaumont et al. 2011; De Beaumont et al. 2012; Miller et al. 2014; Tallus et al. 2012; Tremblay et al. 2011). In addition to their independent use, TMS and EEG can be combined to measure neural activity (with EEG) in response to the TMS pulse. TMS-EEG investigations of brain function outside of the motor cortex are of critical importance given the diffuse nature of mTBI and its cognitive consequences. When applied to the brain, TMS elicits a complex waveform of peaks and troughs, known as a TMS Evoked Potential (TEP), which can be measured using EEG. The TEP is understood to represent the summation of excitatory and inhibitory postsynaptic potentials from large populations of pyramidal neurons (Rogasch and Fitzgerald 2013). EEG captures the propagation of the TMS signal across the scalp, providing information on cortical reactivity, cortical oscillations and local and network connectivity (Rogasch and Fitzgerald 2013). To date, two published studies have measured TMS-EEG outside of the motor cortex in mTBI. These studies reported abnormalities in TEP components in participants an average of 5 years post mTBI (Tallus et al. 2013) and 2 years post mTBI (Opie et al. 2019). Both studies administered TMS to the dorsolateral prefrontal cortex (DLPFC) and neither study evaluated clinical symptom severity or cognitive performance. The research summarised above indicates that TMS-EEG and EEG studies have the potential to provide a greater understanding of the complex relationships between pathophysiology and clinical/cognitive symptoms following mTBI. However, to achieve this, studies need to assess neural activity in cognitively relevant brain regions (at rest, during cognitive processing and in response to external stimulation [i.e. TMS-EEG]), as well as clinical and cognitive symptoms, and the associations between these symptoms and the neural activity. This was the aim of the current study.

We compared individuals who had sustained a mTBI in the previous 4 weeks with individuals with no history of head injury on a comprehensive battery of clinical, cognitive and cortical activity measures. In addition to examining group differences in neuropsychological and neurophysiological measures, we also aimed to explore the relationships between these measures, to assess for interactions between symptoms and pathophysiology. We hypothesised that mTBI participants would report greater clinical symptoms compared to controls and show impairments on cognitive measures that are sensitive to effects of mTBI (e.g. attention, working memory, executive function and processing speed). We also predicted that mTBI participants will exhibit neurophysiological differences on measures of connectivity, functional activity and cortical reactivity (TEP’s) as assessed using resting EEG, task-related EEG and TMS-EEG respectively. The directionality of cortical activity responses were not hypothesised due to the small number of prior studies and heterogeneity in the literature

## 2. Material and Methods

### 2.1 Participants

A total of 58 participants were recruited (30 mTBI, 28 healthy controls). The 30 participants with mTBI were recruited from the emergency department and trauma wards of the Alfred Hospital, Melbourne (less than 1 month post injury, mean days since injury = 19.70, *SD* = 16.96, range 10-31, mean age at injury = 35.43 years, *SD* = 10.31). An age and sex-matched control group was also recruited. These 28 participants had no history of any traumatic brain injury (mean age = 31.65 years, *SD* = 9.06). No participants had a history of seizures, psychiatric or neurological illnesses, unstable medical conditions, were pregnant or prescribed medication known to directly or significantly influence electroencephalographic (EEG) findings. mTBI was classified as GCS 13-15, loss of consciousness < 30 minutes and post-traumatic amnesia (PTA) < 24 hours (Carroll et al. 2004). All participants provided written informed consent prior to commencement of study procedures. The study received approval from both The Alfred and Monash University Ethics Committees.

### 2.2 Procedure

Each participant attended a single 3-hour experimental session where clinical, cognitive and cortical activity measures were assessed. Participants then underwent a session of intermittent Theta Burst Stimulation (iTBS) after which the cortical activity measures were repeated. This was performed to assess the potential modulation of cortical activity by iTBS, results of which will be reported in a separate study. Please note, the current manuscript will report the pre iTBS measures only. All measures are described in detail below.

### 2.3 Clinical and Cognitive Measures

Demographic information included age, date of birth, gender and years of education. The Mini International Neuropsychiatric Interview-Screen (Sheehan et al. 1998) was completed to rule out the presence of psychiatric or substance use disorders. The Edinburgh Handedness Inventory (Oldfield 1971) determined handedness. A range of clinical measures to assess mood, fatigue and post concussive symptomology were administered (see Supplementary Materials for details). Participants completed a comprehensive cognitive battery prior to the neurophysiological assessment. Measures administered included; Wechsler Test of Adult Reading (WTAR), Trail Making Test Parts A and B (TMT), subtests from the Wechsler Adult Intelligence Scale-IV (Working Memory Index and Processing Speed Index), Rey Auditory Verbal Learning Test (RAVLT), Brief Visual Memory Test (BVMT) and Controlled Word Association Test.

### 2.4 Cortical Activity Measures

#### 2.4.1 EEG Recording and Tasks

EEG was recorded with TMS-compatible Ag/AgCl electrodes and a DC coupled amplifier (SynAmps2, EDIT Compumedics Neuroscan, Texas, USA). Fifty electrodes were used from a 64-channel Easycap EEG cap (AF3, AF4, F7, F5, F3, F1, Fz, F2, F4, F6, F8, FC5, FC3, FC1, FCz, FC2, FC4, FC6, T7, C5, C3, C1, Cz, C2, C4, C6, T8, CP5, CP3, CP1, CP2, CP4, CP6, P7, P5, P3, P1, Pz, P2, P4, P6, P8, PO7, PO3, POz, PO8, PO4, O1, Oz, O2). Electrodes were referenced on-line to CPz and grounded to FPz. All data was recorded with a high acquisition rate (10,000Hz) and low-pass filtered (DC-2,000 Hz) using a large operating window (200 mV). Electrode impedances were below 5 kΩ at the start of each resting or task related EEG recording. EEG was recorded at rest and during a number of cognitive tasks (described below). Tasks were presented during EEG using Presentation® software (Version 18.0, Neurobehavioral Systems, Inc., Berkeley, CA, www.neurobs.com), as were instructions provided to participants prior to the resting EEG recordings. EEG pre-processing details are provided in the Supplementary Materials.

#### 2.4.2 Resting EEG

Participants sat quietly and relaxed in a darkened room during resting EEG recordings. Intra-auricular earphones were inserted, and participants instructed to keep their eyes open until they heard a brief tone, after which they should shut their eyes. Recordings of eyes open and eyes closed EEG activity were both 3 minutes in duration.

#### 2.4.3 Working Memory EEG Task - Digit Span Backwards

Participants completed a computerized version of a backwards digit span task. Pseudo-randomized sequential auditory digits were presented aurally through earphones in a performance-adapted list length adjustment design. The stimuli consisted of the digits ‘1’ through ‘9’ and digits could not be repeated directly in any sequence. The task began with digit sequences consisting of 2 digits and the number of digits in each sequence increased by 1 digit every two trials. The task stopped when the participant had two consecutive errors at any given digit set size. A fixation cross was presented for 1000ms to direct participant’s attention, followed by an interval of 1000ms before the presentation of the first digit. Each digit was presented for 500ms with a 1000ms interval. Following presentation of all digits a recall prompt instructed participants to type their response using a keyboard and to press enter to record their response. To calculate participants’ raw digit span score, one point was given for each successful trial and the total points were summed.

#### 2.4.4 Sustained Attention EEG Task - Continuous Performance Test (CPT)

Participants were required to respond with a left mouse click to target (‘Go’) stimuli and inhibit their response to non-target (‘No Go’) stimuli (Riccio et al. 2002). Letters were presented centrally on a computer screen for a time of 300ms (inter stimulus interval of 900ms) in a pseudo randomised order. ‘Go’ stimuli were the letter ‘X’ and ‘No Go’ stimuli were the letter ‘A’. The stimulus set consisted of 244 trials, 50% ‘target’ stimuli, 50% no go stimuli. 10 practice trials were completed prior to task commencing and during the task 30 trials were presented before a rest break was offered. ‘Go’ and ‘No Go’ stimuli were equally probable, so the comparison of brain responses between ‘target’ and ‘non-target’ stimuli was not confounded by the effect of differences in frequency of stimuli (Lavric et al. 2004).

#### 2.4.5 TMS-EEG

TMS was delivered using a figure-of-eight MagVenture B-65 fluid-cooled coil (MagVenture A/S, Denmark) in a biphasic mode. The EEG cap was applied first, and then the resting motor threshold (RMT) was determined (by applying TMS over the cap) as the minimum stimulus intensity required to elicit at least three out of five motor evoked potentials (MEPs) > 0.05 mV in amplitude (Conforto et al. 2004) in the relaxed first dorsal interosseous muscles. TMS was administered to the left dorsolateral prefrontal cortex (DLPFC) at the F3 electrode using the 10/20 system of placement. Participants listened to white noise through intra-auricular earphones (Etymotic Research, ER3-14A, USA) to limit the influence of auditory processing of the TMS click (Rogasch et al. 2014). The sound level was adjusted for each participant until background noise was barely audible. Participants received 100 single pulses at an interval of 4 seconds (with a 10% jitter) at RMT 110%.

### 2.5 Analyses

All statistical analyses of clinical and cognitive measures were performed using R Studio (version 1.1.463) (R Core Team 2018). Resting EEG and TMS-EEG data were processed and analysed offline using EEGLAB (Delorme and Makeig 2004), TMS-EEG Signal Analyser (TESA) (Rogasch et al. 2017), FieldTrip (Oostenveld et al. 2011), BrainNet Viewer (Xia et al. 2013) and custom scripts on the MATLAB platform (version R2017a). EEG task-based data was analysed using Randomisation Graphical User Interface (RAGU), an open-source MATLAB based toolbox (Koenig et al. 2011). Data from two control participants were excluded due to equipment malfunction and experimenter error.

#### 2.5.1 Clinical and Cognitive Measures

Welch’s t-tests were used to evaluate clinical differences between the two groups as they are more robust to unequal variances and unequal sample sizes (Ruxton 2006). Cohen’s *d* was used to calculate effect sizes. One-way ANCOVAs were used to evaluate cognitive differences between the two groups, controlling for premorbid intelligence (WTAR). Assumptions of normality, independence of the covariate and homogeneity of regression slopes were met.

#### 2.5.2 Cortical Activity Measures

##### 2.5.2.1 Resting EEG Data

Following pre-processing (see Supplementary Materials) all participants had provided 60 or more noise free epochs of two seconds in length for eyes open (EO) and eyes closed (EC) conditions, providing enough trials for reliable analysis (EO mean total epochs = 79.95, *SD* = 8.70, EC mean total epochs = 87.68, SD = 6.84). For power computation, EEG data were submitted to a frequency transformation based on fast fourier transform using the ‘mtmfft’ method and Hanning taper (from 0.1 Hz to 100 Hz in steps of 0.2 Hz) to calculate the average power within four frequency bands: theta (4–8 Hz), alpha (8–12 Hz), beta (12–30 Hz) and gamma (30–45 Hz). Average total power was then calculated across all epochs within each frequency band for each condition, resulting in a single value for each participant, within each frequency band and each condition at each electrode. For statistical analysis of power, non-parametric cluster-based permutation statistics assessed differences in power between groups for each frequency band. Conducted in Fieldtrip (Oostenveld et al. 2011), cluster-based permutation tests provide a method for effectively controlling for multiple comparisons across numerous EEG electrodes and time-points (Maris and Oostenveld 2007). Clusters were defined as ≥ 2 neighbouring electrodes with *p* < .05. Monte Carlo p-values were computed on 5000 random permutations. A primary critical α level set at *p* < .05 was used as the cluster-statistical significance for all analyses, controlling for multiple comparisons across space and time, with a secondary threshold for family-wise cluster based null hypothesis testing (*p* < .025; two-tailed test). For connectivity computation, the debiased estimator of the weighted phase lag index (wPLI) was used to compute connectivity from the fourier output of the ‘mtmfft’ frequency measure. The wPLI is conservative measure of phase synchronisation between electrodes. wPLI provides a value for each pair of electrodes between 0 – 1, with higher values reflecting more connectivity between the two electrodes. For statistical analysis of connectivity, the wPLI values for each participant from both groups were averaged across time, epochs, and frequency, in the same frequency bands as power computations for both conditions (EO and EC). Network based statistics (NBS) (a non-parametric statistical method used to make statistical comparisons between large numbers of pairs using cluster based statistical methods to control for multiple comparisons) (Zalesky et al. 2010), was used to analyse differences between groups in each frequency band and each condition. Pairs with a test statistic exceeding the primary threshold (see section 2.4) provide the pairs for the cluster based null hypothesis test. BrainNet Viewer (Xia et al. 2013) was used to visualise significant connections, displaying graph theoretical networks as ball-and-stick models (see Supplementary Materials for additional details on wPLI and NBS methods).

##### 2.5.2.2 Working Memory EEG Data

Following pre-processing (see Supplementary Materials) participants were included only if they had provided =>15 artefact-free correct epochs of 1 second in length. The final sample included 26 mTBI participants (Number of epochs; *M* = 38.78, *SD* = 17.35) and 23 control participants (Number of epochs; *M* = 31.27, *SD* = 13.73). Only individuals included in the EEG analysis were included in the digit span backwards behavioural analyses. For statistical analysis of ERP’s, measures of neural response strength and topography were calculated using RAGU with 5000 randomization runs, and a threshold of *p <* .05 (see Supplementary Materials).

##### 2.5.2.3 CPT EEG Data

Following pre-processing (see Supplementary Materials) participants were included if they had a minimum of 80 artefact-free correct epochs of 1 second in length for ‘Go’ and ‘No Go’ conditions. The sustained attention task was added to the protocol as an additional measure after recruitment had commenced, resulting in a smaller sample size. The final sample included a total of 17 control participants (Go-epochs: *M* =□112.70, *SD* = 4.59; No Go-epochs: *M* =□107.41, *SD* = 8.80) and 20 mTBI participants (Go-epochs: *M* =□111.10, *SD*L = □5.78; No Go-epochs: *M* =□107, SD = □8.16). To assess whether behavioural performance differed between control and mTBI participants, separate two-way ANCOVAs were conducted for accuracy and reaction time, with group (control vs. mTBI), condition (‘Go’ vs ‘No Go’) and pre-morbid IQ (WTAR). For statistical analysis of ERP’s, measures of neural response strength (global field power, GFP) and topography were calculated using RAGU (Koenig et al. 2011) (see Supplementary Materials).

##### 2.5.2.4 TMS-EEG Analyses

Following pre-processing (see Supplementary Materials) all participants provided a minimum of 55 artefact free epochs of 2 seconds in length. One mTBI participant was removed from the TMS-EEG analysis due to problems with the recording file. The final sample included 29 mTBI (Number of epochs; *M* = 94.3, *SD* = 7.48) and 26 control participants (Number of epochs; *M* = 97.7, *SD* = 3.05).

TEP analysis focussed on four separate peaks known to occur following stimulation of the prefrontal cortex, the N45, P60, N100 and P200 (Rogasch et al. 2015; Rogasch et al. 2014). Negative peaks were identified as the maximum occurring peaks between 0.035 and 0.050 s following the TMS pulse (N45), and 0.09 and 0.135 s (N100); while the positive peaks were identified as those occurring between 0.05 and 0.07 s (P60) and 0.15 and 0.24 s (P200). Cluster based analyses, as per resting EEG data, were performed to test for differences between groups (Control vs. mTBI) using t-test designs. For significant clusters, regions of interest were defined, and peak amplitude values extracted for TEP components.

#### 2.5.3 Correlations

To assess relationships between clinical, cognitive and cortical activity measures Pearson’s correlations were used. To reduce the number of comparisons, associations were only explored between measures that significantly differentiated the groups.

## 3. Results

Demographics for the mTBI and control participants are summarised in Table 1. The groups did not differ significantly on measures of sex, age and pre-morbid intelligence (all p > .05), however controls had a higher level of education (p = .035).

**Table 1.** Demographic Information for Control and mTBI

| | Control | mTBI | | $p$ | $d$ |
| --- | --- | --- | --- | --- | --- |
| $N$ | 26 | 30 | | | |
| sex = male (%) | 18 (69.2) | 23 (76.7) | $\chi^2 = .11$ | 0.746 | 0.087 |
| age (mean (sd)) | 31.65 (9.06) | 35.43 (10.31) | $t = -1.46$ | 0.154 | 0.389 |
| education (mean (sd)) | 16.94 (2.59) | 15.25 (3.19) | $t = 2.19$ | 0.035* | 0.582 |
| WTAR (mean (sd)) | 41.76 (5.63) | 38.00 (7.86) | $t = 1.91$ | 0.053 | 0.506 |

### 3.1 Clinical Measures

All individuals received a CT scan upon their admission to the Alfred Hospital, presence of haematoma, haemorrhage or fracture was classified as CT pathology positive in line with previous classification of “complicated” vs “uncomplicated” mTBI (Williams et al. 1990). Clinical characteristics of the mTBI group are presented in Table 2. “Other injury” was classified as orthopaedic or musculoskeletal injuries co-occurring at the time of mTBI.

**Table 2.** Clinical characteristics of mTBI group

| Variables | $N = 30$ |
| --- | --- |
| Days post injury (M/SD/range)) | 19.70/ 16.96/ 10-31 |
| Short (0-10 days post) | 6.7% |
| Medium (11-20 days post) | 46.6% |
| Long (21-31 days post) | 46.6% |
| Amnesia |  |
| Retrograde | 33.3% |
| None | 30% |
| Retrograde and anterograde | 23.3% |
| Unknown | 13% |
| GCS |  |
| 15 | 53.3% |
| 14 | 20% |
| 13 | 3% |
| unknown | 23.3% |
| LOC (%) |  |
| Yes | 40% |
| No | 43.3% |
| Unknown | 16.7% |
| PCS severity |  |
| Mild | 30% |
| Moderate | 36.7% |
| Severe | 10% |
| None | 23.3% |
| History of mTBI (%) |  |
| Yes | 56.7% |
| No | 43.3% |
| CT pathology (%) |  |
| Yes | 43.3% |
| haematoma | 23.3% |
| haemorrhage | 6.7% |
| haematoma & haemorrhage | 6.7% |
| haematoma & fracture | 6.7% |
| No | 53.3% |
| Unknown | 3.3% |
| Mechanism of Injury (%) |  |
| Fall from bicycle | 43.3% |
| Fall | 20% |
| Head strike by object | 13.3% |
| Pedestrian vs. motor vehicle | 13.3% |
| Motor vehicle accident | 9.9% |
| Other injury at time of accident |  |
| Yes | 63.3% |
| No | 36.7% |

Measures for mood and symptom evaluation for control and mTBI participants are presented in Fig 1. The mTBI group showed significantly greater scores on measures of anxiety, depression, fatigue and post-concussion symptoms (*p* < .05). 23.3% (n=7) reported anxiety symptoms in the elevated range and 3.33% (n=1) in the clinically significant range. For depressive symptoms, 16.7% (n=5) of mTBI participants were in the elevated range and 6.67% (n=2) in the clinically significant range. See Supplementary Table 2 for means and standard deviation.

**Fig 1:**
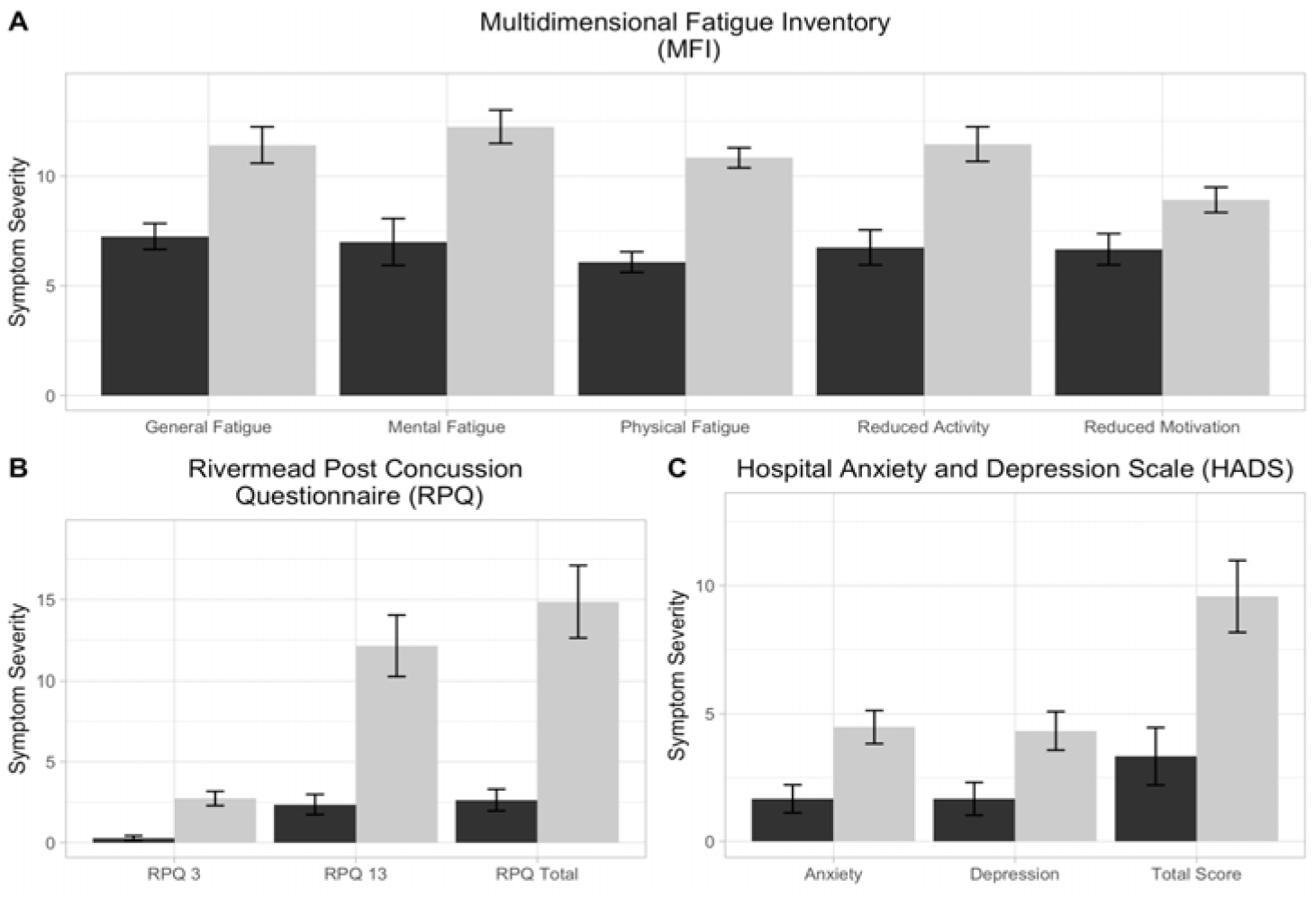
Clinical symptoms for mild traumatic brain injury (mTBI) and control group participants. This figure shows participant’s symptom report on A) fatigue (measured by the Multidimensional Fatigue Inventory (MFI)), B) mood (measured by the Hospital Anxiety and Depression Scale (HADS)) and C) post-concussion symptom (measured by the Rivermead Post Concussion Symptom Questionnaire (RPQ)) measures. Control participants are indicated in *black* and mild traumatic brain injury (mTBI) participants in *grey*. Values depicted are means with standard error bars. See supplementary information (Table S2) for additional information. P values correspond to the outcome of Welch’s t-tests and all comparisons yielded *p* < .05.

### 3.2 Cognitive Measures

Performance on cognitive tasks for control and mTBI participants are presented in Table 3. To account for the higher level of education in the control group, co-variate analyses were conducted for cognitive measures controlling for pre-morbid intelligence (WTAR). This measure was chosen instead of education as it did not violate the ANCOVA independence of covariate and dependent variable assumption. ANCOVA demonstrated that mTBI participants recalled fewer words on the first trial of the RAVLT, *F* (1, 51) = 6.13, *p* = 0.02, η_p_^2^ = 0.11 and reached trend significance on delayed recognition *F* (1, 48) = 3.92, *p* = 0.05, η_p_^2^ = 0.08. No other significant differences in cognitive performance on pencil and paper measures were shown.

**Table 3.** Descriptive statistics and ANCOVA results for cognitive measures

|  | Control | mTBI |  |  |
| --- | --- | --- | --- | --- |
|  | M (SD) | M(SD) | *F* | *p* |
| *N* | 26 | 30 |  |  |
| Trail Making Test(seconds) |  |  |  |  |
| TMT A | 25.71 (9.14) | 26.69 (9.10) | 0.03 | 0.86 |
| TMT B | 50.29 (16.40) | 57.94 (20.61) | 0.02 | 0.95 |
| RAVLT |  |  |  |  |
| Trial 1 | 8.04 (1.95) | 7.00 (1.64) | 6.13 | 0.02* |
| Trial 2 | 10.54 (2.10) | 9.73 (2.38) | 0.94 | 0.33 |
| Trial 3 | 12.46 (2.12) | 11.27 (2.00) | 3.23 | 0.07 |
| Trial 4 | 12.81 (1.88) | 11.97 (1.77) | 1.43 | 0.24 |
| Trial 5 | 13.50 (1.53) | 13.03 (1.67) | 0.30 | 0.58 |
| Total (T1-T5) | 57.12 (8.14) | 52.90 (8.13) | 2.43 | 0.13 |
| Delayed Recall | 12.23 (2.14) | 11.10 (2.99) | 2.39 | 0.12 |
| Delayed Recognition | 40.12 (4.19) | 37.59 (4.13) | 3.92 | 0.05 |
| BVMT-R |  |  |  |  |
| Total (T1-T3) | 27.38 (4.65) | 28.93 (3.99) | 3.58 | 0.06 |
| Delay | 10.81 (1.10) | 10.96 (1.43) | 0.23 | 0.64 |
| Digit Span Forward | 11.50 (2.90) | 10.80 (2.41) | 0.01 | 0.93 |
| Digit Span Backward | 8.50 (2.60) | 7.33 (1.81) | 1.29 | 0.26 |
| Arithmetic | 15.35 (3.57) | 15.62 (4.04) | 2.63 | 0.11 |
| Letter Number Sequencing | 11.96 (2.58) | 11.52 (2.82) | 0.26 | 0.61 |
| Coding | 63.96 (11.32) | 58.17 (7.68) | 2.43 | 0.12 |
| Symbol Search | 36.92 (6.88) | 35.40 (6.99) | 0.35 | 0.56 |

$p < 0.05$

### 3.3 Cortical Activity Measures

#### 3.3.1 Resting EEG Data

##### 3.3.1.1 Power Analysis

mTBI participants were shown to have greater alpha power in the right fronto-central region during the eyes closed condition (t = -30.02, p = 0.0154) (see Fig 2). No significant differences were demonstrated in theta, beta or gamma frequencies (p > .05).

**Fig 2.**
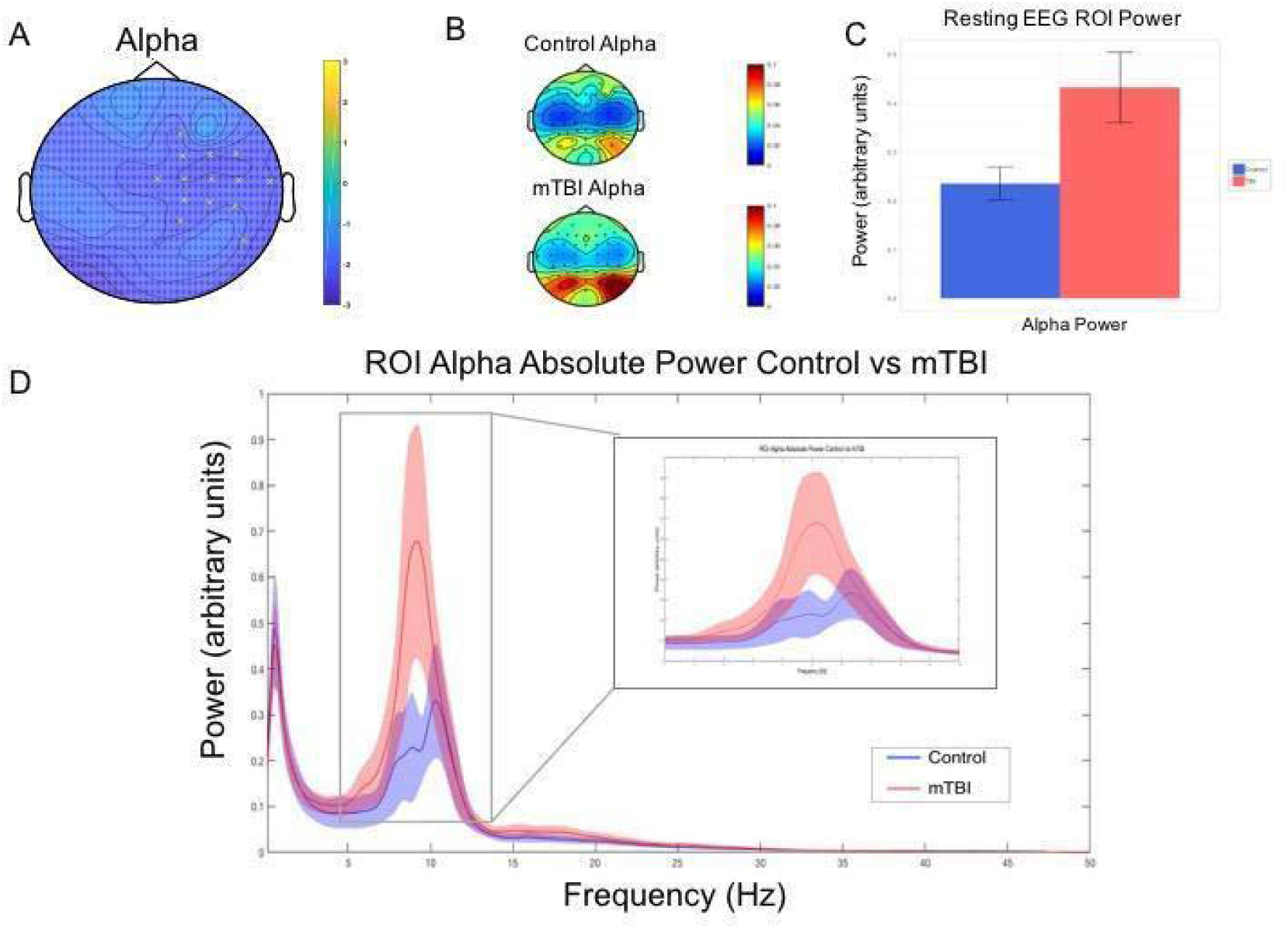
Resting EEG alpha power group differences. This figure shows mild traumatic brain injury (mTBI) participants demonstrated significantly greater alpha power (8–12 Hz arbitrary units) during the eyes closed resting EEG condition compared to the control group. A— Topoplot showing adjacent electrodes (*p* < 0.025) clustering in in the right fronto-central region when testing for a difference in power in the alpha range (8-12 Hz) between groups. B— Topoplot showing the distribution of activity between groups for the region of interest (ROI) as identified by the clustering electrodes. C—Alpha power for ROI electrodes for control and mTBI participants (values depicted are means with standard error bars). D— Mean spectral power content from cluster ROI electrodes with 95% confidence interval shading.

##### 3.3.1.2 Connectivity Measures (wPLI)

Using the wPLI connectivity matrices for all electrode pairs, comparisons between control and mTBI participants demonstrated no significant differences in connectivity across theta (4-8Hz), alpha, (8-12hz), beta (12-30hz) and gamma (30-45Hz) bands between the two groups (*p* > .05) across eyes open and eyes closed conditions.

#### 3.3.2 Working Memory EEG Data

##### 3.3.2.1 Task related EEG

The GFP test showed a significant main effect of group from 617-655ms and 950-1000ms post digit presentation (exceeding the global duration control of 25 ms). However, these periods were during periods of topographical inconsistency in the TCT test, suggesting that any group differences during these periods may be the result of inconsistent neural activity patterns within groups, rather than a significant difference between the groups. As such, these differences were not interpreted further. The TANOVA demonstrated no significant topographical differences in the distribution of neural activity (see Supplementary Materials).

##### 3.3.2.2 Behavioural Data

Differences in digit span performance accuracy were subjected to a Welch’s independent samples t-test. There was no significant effect of group, with the control group recalling a non-significantly higher number of items on the digit span task (*M =* 9.61, *SD =* 2.43) than the mTBI group (*M =* 8.50, *SD =* 2.57*), t* (47) = 1.55, *p* = 0.127.

#### 3.3.3 CPT EEG Data

##### 3.3.3.1 Task Related EEG

The 2 × 2 (Group x Condition) global field power (GFP) test showed a significant main effect of condition in a period from 376-428 ms post stimuli presentation (global count *p* = 0.0062, which exceeded the global duration control of 38 ms). Average GFP was significantly higher in response to the ‘Go’ compared to the ‘NoGo’ condition. No main effect of group or significant between group and condition interaction was demonstrated in the GFP test (*p* > .05). The TANOVA demonstrated no significant topographical differences in the distribution of neural activity for group at any point across the epoch (*p* > .05). A main effect of condition was demonstrated, see Supplementary Materials.

##### 3.3.3.2 Behavioural Data

Differences in sustained attention task performance (reaction time and accuracy) were subjected to separate two-way ANCOVA’s. For reaction time, there was a significant effect of condition on reaction time after controlling for pre-morbid IQ, participants responding significantly faster to ‘No Go’ trials (*M* = 258.23, *SD* = 33.31) than ‘Go’ trials (*M* = 318.50, *SD* = 30.75), *F* (1,64) = 54.51, *p <* 0.01. For accuracy, a significant effect of condition after controlling for premorbid IQ was also demonstrated, control and mTBI participants responding more accurately to ‘Go’ trials (*M* = 0.997, *SD* = 0.004) than ‘No Go’ trials (*M* = 0.952, *SD* = 0.050), *F* (1, 67) = 29.39, *p* < 0.01). For reaction time and accuracy, no significant effect of group or significant interaction between group and condition was demonstrated (*p* > .05).

#### 3.3.4 TMS-EEG Data

Single-pulse TMS over the left dorsolateral prefrontal cortex (DLPFC) resulted in a characteristic series of negative and positive peaks including N45, P60, N100 and P200. The grand average TEP for each group is displayed in Fig 3, with the four peaks of interest (N45, P60, N100, P200) and their respective time windows outlined.

**Fig 3.**
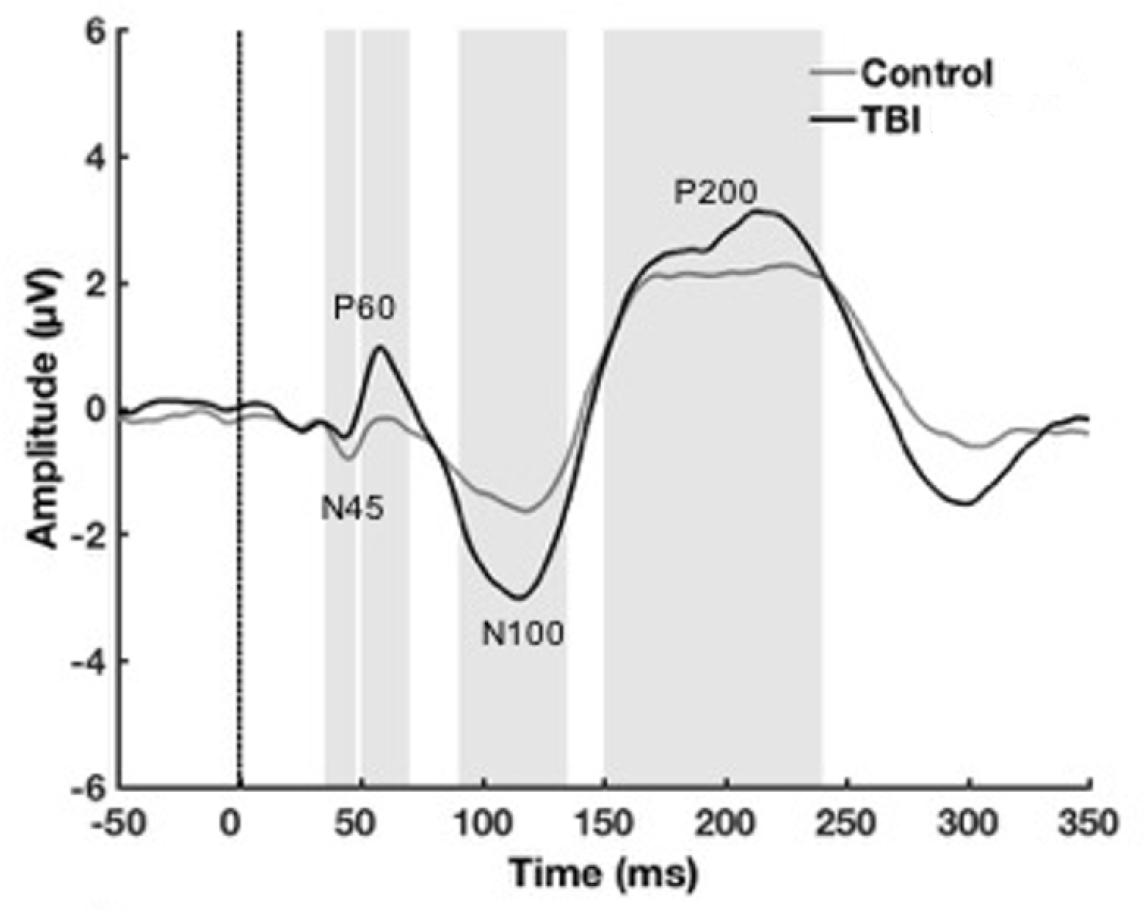
Transcranial-evoked potential (TEP) grand average waveforms. This figure plots TEP grand average waveforms following delivery of TMS pulse (shown with dotted line) across all electrodes for each group. Peak windows of interest (N45, P60, N100 and P200) that underwent analysis are delineated by grey shading.

Non-parametric cluster-based permutation tests revealed a significant difference between groups for the P60 TEP that was most pronounced in the left parieto-occipital region (*t* = 23.10, *p* = 0.012) (Fig 4). For the N100 TEP component there was a significant between group difference that was most pronounced in the right fronto-central region (*t* = 36.26, *p =* 0.0036) and in the left parieto-occipital region (*t* = - 31.66, *p =* 0.0058).

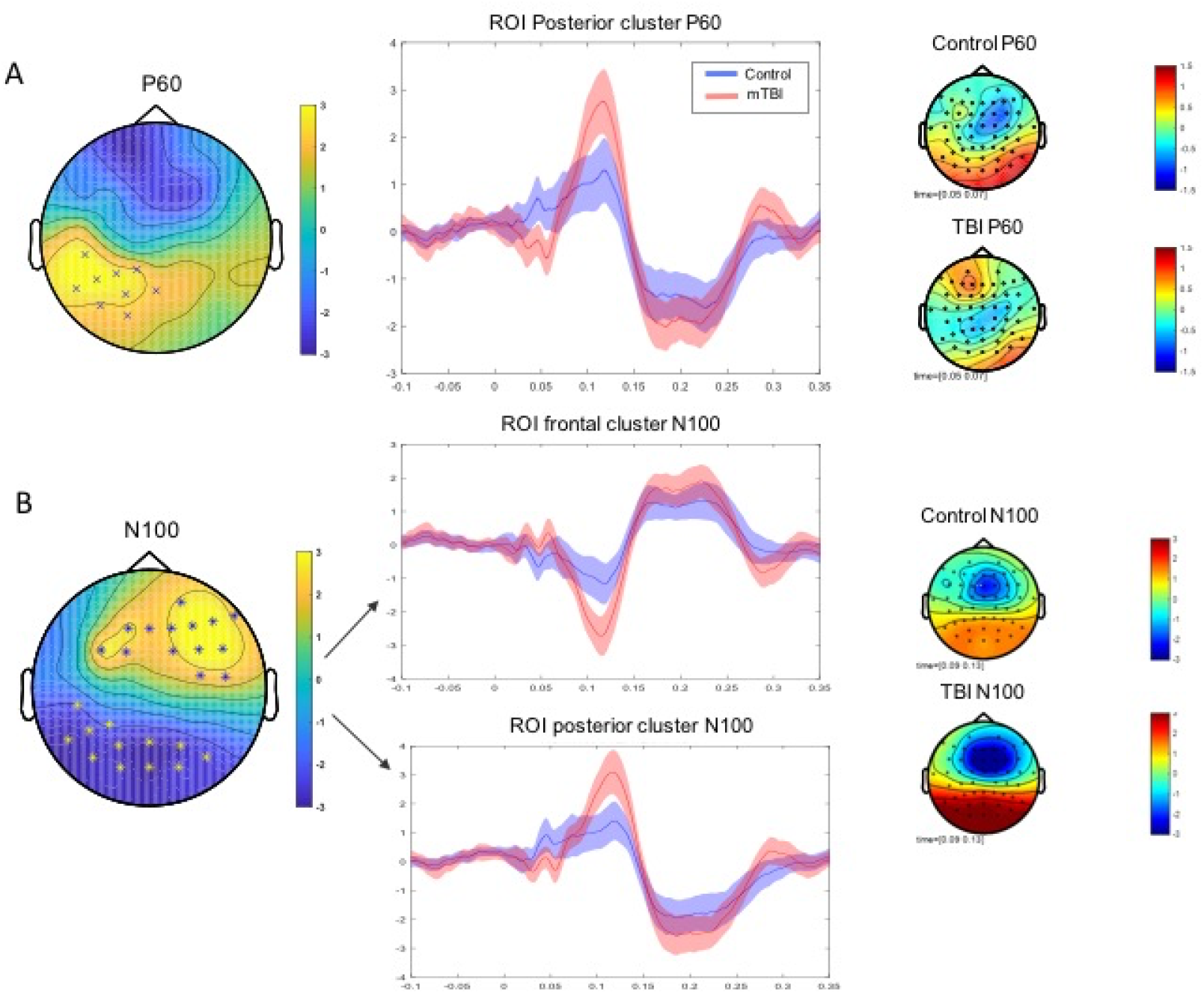

Electrodes identified as forming clusters when testing for the N100 and P60 TEP measures were used to develop regions of interest (ROI). ROI peak to peak mean amplitude was extracted (see Fig 5) to be used to explore relationships with clinical and cognitive measures.

**Fig 5.**
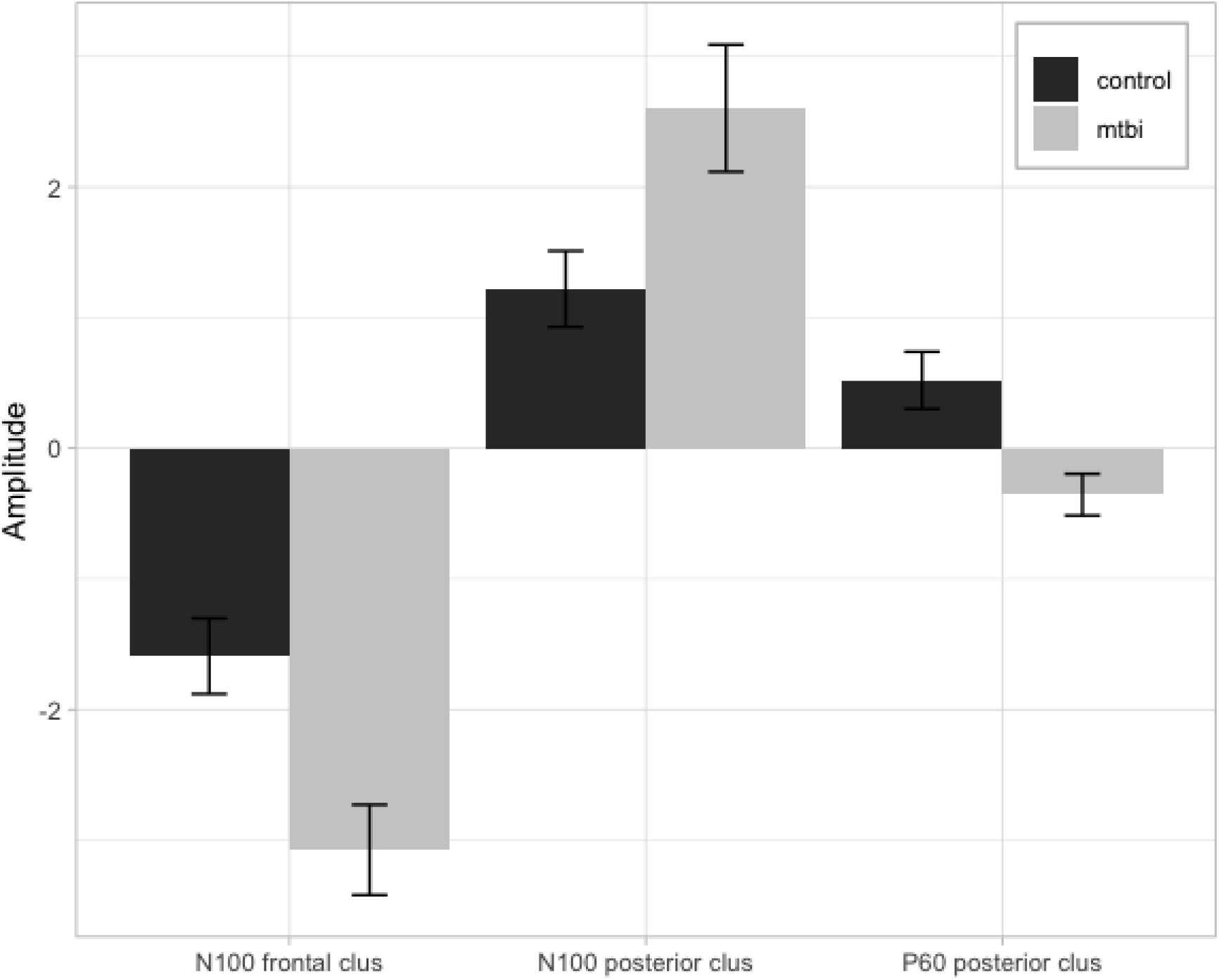
Peak to peak amplitude. This figure plots transcranial evoked potential (TEP) peak to peak mean amplitude values and standard error for region of interest (ROI) analyses for N100 (frontal and posterior) and P60 (frontal) TEP components in control (black) and mTBI (grey) groups (as indicated in Fig 3).

### 3.4 Correlations

Associations between behavioural measures and neurophysiological markers of pathophysiology that significantly differentiated the groups were assessed using correlational analyses. Clinical and cognitive measures included; mood, fatigue, PCS severity and verbal learning. Cortical activity measures included; resting alpha power and P60 and N100 TEP amplitude. No significant associations were demonstrated between resting EEG alpha power and clinical or cognitive measures in either group (*p* > .05). For TEP measures, no associations between posterior or anterior N100 amplitude or posterior P60 amplitude and clinical or cognitive measures were demonstrated (*p >* .05).

## 4. Discussion

The current study comprehensively characterised the clinical, cognitive and neural impact of mTBI. As expected, mTBI participants reported higher rates of post-concussive, depressive, anxious and fatigue symptoms than controls. Cognitively, on a measure of verbal learning, mTBI participant performance was significantly below that of age and gender matched controls, after controlling for premorbid IQ. Cortical activity changes were investigated with a variety of methods, including resting and task-based EEG and TMS-EEG. Overall, mTBI participants demonstrated increased alpha power at rest and altered cortical reactivity as measured via TMS-EEG. There were no significant correlations found between pathophysiology and the clinical/cognitive symptoms identified.

There are conflicting reports in the previous literature on mTBI of the presentation and persistence of symptoms and cognitive impairment. Our results support previous literature that argues the impact of mTBI is not inconsequential (McInnes et al. 2017). mTBI participants reported significantly greater symptoms on mood, fatigue and PCS measures compared to controls when assessed in the subacute phase. A history of pre-morbid psychopathology was carefully screened for and excluded, suggesting mood symptoms were not associated with pre-existing psychiatric factors. However, levels of anxiety and depression symptoms were not, on average, in the clinically significant range. Cognitively, the mTBI group showed reduced performance on some measures of verbal learning. Verbal learning deficits (as measured by reduced total immediate learning and delayed recall) are associated with moderate-severe TBI (Draper and Ponsford 2008) but have been less consistently demonstrated in mTBI (Albrecht et al. 2016; Ettenhofer and Abeles 2009). It has been suggested that the choice of verbal learning metric may mediate findings, with total immediate learning and delayed recall being insufficient to detect subtle deficits (Bigler 2008). Our mTBI group demonstrated impaired initial acquisition (i.e. the first learning trial) but comparable total recall, suggesting a slower rate of learning but equivalent performance across time. This may be due to reduced usage of efficient internally driven strategies that facilitate learning (Geary et al. 2010). Learning and memory are closely linked with attention, working memory and executive functioning (McCabe et al. 2010), suggesting reduced initial acquisition may be mediated by fluctuating attention, reduced planning and strategy generation in mTBI participants. Impaired delayed recognition performance reached trend significance and is likely to have been mediated by similar deficits, with reduced initial acquisition on an interference word list, impacting overall recognition performance of both lists on delay. Commensurate performance on other cognitive measures, but increased fatigue, may reflect increased effort to maintain task performance in our mTBI group. Having established group level differences our study next looked to characterise relevant cortical activity changes.

Our mTBI group demonstrated greater right fronto-central alpha power during an eyes closed condition compared to control participants. This finding is somewhat inconsistent with previous literature, with greater delta and theta and reduced alpha power having been previously reported. However, the previous literature differs from our study - one study reported mTBI had occurred in the past 12 months (Gosselin et al. 2009), one used a military mTBI sample (Lewine et al. 2019) and another has been criticised for misclassifying moderate to severe injuries as mTBI (Korn et al. 2005). The dynamic nature of mTBI, neural changes evolving across recovery (Coyle et al. 2018; Eierud et al. 2014) and variability in findings based on injury severity, suggest our results may not be comparable due to divergent samples. Interpretation of our findings is further complicated by controversy regarding the functional significance of alpha power. Resting and task-related alpha has been associated with contradictory physiological and cognitive processes. The dominant theory links alpha oscillations to inhibitory function (Jensen and Mazaheri 2010), however, these conclusions have been drawn from studies predominantly focused on cognitive tasks (Jensen and Mazaheri 2010; Palva and Palva 2007; Sadaghiani et al. 2012). Physiologically, alpha activity originates from thalamo-cortical neurons (Lorincz et al. 2009) by means of GABA-ergic interneurons (Hughes et al. 2011). Resting state thalamo-cortical networks have been shown to be disrupted in mTBI (Tang et al. 2011) and thalamic damage has been hypothesised to contribute to impairments post mTBI (Grossman and Inglese 2016). As such, increased right fronto-central alpha power may be related to the underlying pathophysiology of mTBI, however further investigations are needed. Network based analyses, that investigate inter regional connectivity, are of relevance due to the diffuse nature of mTBI. Long-distance connectivity has been shown to be decreased and short-distance connectivity increased, suggesting changes to the functional architecture following mTBI (Cao and Slobounov 2010). Increases in connectivity in the frontal areas of the brain are hypothesised to reflect mechanisms compensating for structural connectivity losses (Palacios et al. 2013b) or increased effort in recruiting the appropriate neural networks (Caeyenberghs et al. 2014). We used wPLI, a synchronization measure, to assess whole brain connectivity at rest. No differences between groups were demonstrated in the theta, alpha, beta or gamma bands. This is inconsistent with reported decreases in low-gamma frequency band (25-40Hz) (Wang et al. 2017) or increased gamma activity (30-45Hz) during a working memory task (Bailey et al. 2017) in mTBI. Although our sample is comparable, differences in methodology may account for our differential findings. wPLI is a conservative measure of connectivity, and although it controls for false positives from volume conduction, it also may have failed to identify true differences between the groups by being overly rigorous.

We also recorded EEG during cognitive tasks. No differences were detected between the mTBI group and control group in a backwards digit span working memory task. This is not consistent with previous research demonstrating EEG abnormalities during working memory tasks (Arakaki et al. 2018; Kaltiainen et al. 2019; Theriault et al. 2011). However, variability in task stimuli, cognitive load, method of data quantification and type of EEG measures limit comparisons with previous research. Our measure of neural response strength, global field power (GFP) is widely used in clinical neurophysiology as a parameter of total underlying brain activity. No differences in pattern of activation (topography) were demonstrated. These results suggest that perhaps neural activity related to working memory is not altered in mTBI participants.

We also sought to assess inhibitory control and sustained attention using a continuous performance task (CPT), also known as a Go/No Go task. Our results did not show differences in GFP, a measure of neural response strength, or neural topographies, which is consistent with previous research in a group of mild-moderate TBI using an emotional Go/NoGo task paradigm (Bailey et al. 2014). Our task presented ‘Go’ and ‘No Go’ trials with 50% probability and an inter-stimulus interval of 900 ms, which may have insufficiently taxed response inhibition processes to discriminate between the groups. As a consequence, neither group may have been generating sufficient amplitudes of response inhibition processes for comparison of these processes (Wessel, 2018). The lack of behavioural differences may also be due to a ceiling effect in the behavioural data – as average performance in both groups was over 90% for ‘Go’ and ‘No Go’ stimuli.

The application of TMS over the left DLPFC with concurrent EEG is a promising approach to measuring neural activity in mTBI. Significantly greater right fronto-central N100 and smaller left parieto-occipital P60 and greater N100 amplitudes were demonstrated in mTBI participants compared to controls. TEPs are broadly considered a measure of cortical reactivity, with some evidence that the N100 amplitude may reflect, in part, GABA_B_ mediated inhibition (Premoli et al. 2014). While the P60 component has been shown to be sensitive to changes in excitability (Voineskos et al., 2019). Our N100 finding is consistent with the single known study that has administered TMS outside of the motor cortex in mTBI. Tallus et al. (2013) investigated TEP’s in symptomatic and asymptomatic mTBI participants on average five years since injury. Results demonstrated latency delays for early TEP components and higher N100 amplitudes in symptomatic participants, compared to asymptomatic and controls (Tallus et al. 2013). Changes were interpreted as suggestive of altered brain reactivity and connectivity in mTBI and possibly related to compensatory mechanisms of recovery. The smaller P60 and increased N100 amplitudes seen in mTBI suggest changes to synaptic excitation and inhibition, and the balance between the two, with likely consequences for neuronal signalling and communication.

However, it is currently unclear whether these changes reflect ongoing pathophysiology, compensatory reorganisation or the early stages of recovery. Accurately characterising how the balance of neural circuits underlies symptom recovery may assist in the development of diagnostic and prognostic indicators and improve our understanding of symptom heterogeneity post mTBI. We did not see any significant correlations between the measures of pathophysiology and the persistent clinical/cognitive symptoms identified. There are a number of possible reasons for this including, the analyses being underpowered, narrow inclusion of pathophysiologies and narrow focus on specific clinical/cognitive symptoms. A true lack of any association between pathophysiology and clinical/cognitive symptoms is unlikely, and this question requires further investigation with larger samples and broader inclusion of changes in cortical activity and symptomatology.

Several limitations should be noted. Approximately 50% of mTBI individuals sustained a concurrent injury in addition to their mTBI and 16.7% (n=5) were taking analgesic medication. mTBI participants have been shown to report a similar number of PCS to trauma patients (Landre et al. 2006), suggesting pain, medication or other factors may be mediating their symptom report and cognitive performance. Use of orthopaedic control groups has previously been recommended to help determine whether subjective symptoms are specific to the mTBI. There is some evidence however, that cognitive findings are unrelated to pain intensity or mood disturbances (Carroll et al. 2014). A second limitation is that most participants were assessed in the medium (11-20 days) and long (21-31 days) time windows post injury, a period during which recovery rapidly occurs and symptoms resolve for most individuals with mTBI (McCrea et al. 2009). Therefore, group differences may have been more apparent if assessment had occurred closer to injury time and it is possible that individuals in the short (0-10 days) time window differed in symptoms and pathophysiology. Third, the control group had a higher level of education than the mTBI group and although we included pre-morbid IQ as a covariate, this difference may have mediated cognitive performance. Regarding cortical activity measures, we used the F3 scalp location as an approximate landmark for the DLPFC, over which single pulse TMS was administered. Although this method has been shown to provide a relatively accurate estimate of the DLPFC (Rusjan et al. 2010), future research utilising MRI-based neuro-navigational software could help to further increase accuracy when targeting this region. Lastly, cluster-based permutation tests are useful at identifying significant differences between groups. They do not, however, establish the brain region or time window of an effect, limiting the spatial and temporal inferences of reported differences.

The present study comprehensively characterised the multidimensional effect of mTBI in the sub-acute phase post injury, a broad range of clinical, cognitive and cortical activity consequences being reported. Collectively, the differences between mTBI and control groups may indicate neurophysiological markers of altered excitatory and inhibitory processes which impact neural signalling. Improved understanding of these factors and how they contribute to symptom heterogeneity could assist with the development of potential therapeutic targets.

## Data Availability

We do not have ethical approval to make this data publicly available, as our approval
predated our inclusion of such approvals (which we now do routinely).

## Funding

This work was supported by an Australian Postgraduate Award Scholarship (HLC) and a National Health and Medical Research Council Fellowship (1135558) (KEH).

## Data and Code Availability Statement

We do not have ethical approval to make this data publicly available, as our approval predated our inclusion of such approvals (which we now do routinely).

## Declaration of Competing Interest

None of the authors have potential conflicts of interest to be disclosed.

## Notes

### Competing Interest Statement

The authors have declared no competing interest.

### Clinical Trial

ACTRN12616001038482

### Author Declarations

Ethics Committees of both Alfred Health and Monash University gave ethical approval for this work.

