## Supplementary Materials for "Clinical symptoms, cognitive performance and cortical activity following mild traumatic brain injury (mTBI)"

### **Materials and Methods**

##### Clinical Measures

Table S1. Description of Clinical Measures

| Measure | Description |
| --- | --- |
| *The Rivermead Post-Concussion Symptom Questionnaire (RPCSQ)* (King et al. 1995) | 16 item self-report questionnaire based on a subjective five-scale rating of 0–4, relative to the premorbid levels, of 16 most commonly reported mTBI symptoms, with 0 indicating not experienced, and 4 severe. Categorical variables were created as follows; nil (0), mild (1-16), moderate (17-32), severe (33-48), very severe (49-64) symptoms. |
| *Rivermead Post-Concussion Symptom Questionnaire- Control Version (RPCSQ-C)* (King et al. 1995) | 16 item self-report questionnaire. This measure is included to assess and control incidence of PCS symptoms in controls with no history of head injury. |
| *Hospital Anxiety and Depression Scale (HADS)* (Zigmond and Snaith 1983) | 14 item self-report questionnaire to evaluate rates of depression, anxiety and emotional distress amongst participants. An anxiety or depression total score of >=7 was considered elevated and >= 10 clinically significant. |
| *Multidimensional Fatigue Inventory (MFI)* (Smets et al. 1995) | 20 item self-report questionnaire to evaluate different aspects of fatigue. Yields five domains (General Fatigue, Physical Fatigue, Reduced Activity, Reduced Motivation, Mental Fatigue) |

##### Analyses

###### Resting EEG Pre-Processing

Resting EEG data were down sampled (500 Hz), bandpass filtered (fourth-order, zero-phase, Butterworth filter, 0.1-100 Hz), bandstop filtered (48-52 Hz; to remove 50Hz line noise) and epoched into two second epochs. Automatic artefact rejection was completed, which first checked if more than 3% of epochs included electrodes that varied by more than -250 to 250 microvolts and excluded those electrodes. Next epochs were excluded if they showed a variation of more than 5 SD’s of kurtosis for individual channels, or 3 SD’s for all channels. Lastly, epochs with power within the frequencies 25 to 45Hz that exceeded -100 or 30 dB were excluded (power in these frequencies usually reflects muscle activity). Manual artefact rejection was then completed to ensure the automatic process did not miss significant artifacts, the data being visually inspected to remove epochs with excessive noise (i.e. muscle artefact), and bad channels (i.e. disconnected). Independent component analysis (Fast-ICA algorithm using the ‘tanh’ constrast function) decomposition were then applied to the data. Rejected electrodes were re-constructed using spherical interpolation (Perrin et al. 1989), and data was re-referenced to the average reference. Resting EEG data was then split into two files (at the marker for the auditory tone), creating separate files for eyes open (EO) and eyes closed (EC) conditions.

###### Resting EEG Connectivity Analyses

For connectivity computation, wPLI measures were chosen because they are robust against the effects of volume conduction, non-brain related artefact and activity from a common reference. Phase lags between sensors of near zero contribute minimally to the wPLI measure, preventing the detection of false positive connectivity due to these artefacts (Vinck et al. 2011). For statistical analysis of connectivity, the network-based statistic approach was chosen. NBS is the graph analogue of cluster-based statistical methods, however rather than clustering in physical space, the NBS clusters in topological space. First, the NBS tests the null hypothesis at every pair of nodes, providing a test statistic value for each pair. Connectivity matrices are inputted, with nodes (which correspond to electrode coordinates) and edges (which correspond to connectivity strength, i.e. wPLI values), allowing control and mTBI participants’ connectivity strength values to be compared across every pair of electrodes (Zalesky et al. 2010). See Zalesky et al. (Zalesky et al. 2010) for further information on this analysis technique.

###### Digit Span EEG Pre Processing

Digit Span EEG data were down sampled (1000 Hz), bandpass filtered (fourth-order, zero-phase, Butterworth filter, 0.1-100 Hz) and bandstop filtered (48-52 Hz; to remove 50 Hz line noise). Because temporal relationships were of interest in the task related EEG data a higher sampling rate was used than in the resting data to improve temporal precision. A custom function appended the Presentation .log file to the EEG data to label participant’s responses as ‘Correct’ and ‘Incorrect’. Digit span EEG files from Pre iTBS and Post iTBS EEG recordings were then merged to provide more datapoints for artifact rejection (as ICA performs better with more data (Korats et al. 2012), but note that the files were later split so only the pre iTBS data was analysed). Automatic and manual artefact rejection was completed as per the resting EEG data, except that AMICA (Adaptive Mixture of Independent Component Analysis) (Palmer et al. 2011) was used to manually select and remove eye movements and remaining muscle activity artefacts (instead of Fast-ICA). To ensure data was in the correct form for RAGU analysis, previously merged Pre and Post files were split, referenced to the average reference, correct responses were baseline corrected to the period from -500 ms to stimulus onset, and epoched from digit presentation to 1000ms post.

Prior to statistical analysis, automatic outlier detection using multidimensional scaling in RAGU was used to detect and exclude extreme values which resulted in the removal of one control and one mTBI participant. The automatic outlier detection is based on an algorithm that uses the Mahalanobis distance among the displayed points to identify cases that are unlikely to be part of the normal distribution (Habermann et al. 2018). Additionally, due to the performance-adapted list length adjustment design nature of the task, the number of epochs per participant varied. Only participants with =>15 correct epochs per time point (Pre and Post) were included in the analysis. This number of epochs was chosen to maximise data inclusions. If a more conservative cut-off (=>30 correct epochs) was applied, only 16 control and 11 mTBI participants would have been included in the analyses. An independent samples t-test was used to compare number of included epochs between groups which detected no significant difference (*t (47) =* 1.691, *p* = 0.098). Total mean number of epochs for the control group (*M* = 38.78, *SD* =17.35) was similar to the mTBI group (*M* = 31.27, *SD* =13.73).

A topographic consistency test (TCT) was then conducted to identify the periods in the epoch which there is positive evidence for a consistent distribution of scalp activity across participants within each group and time point (Koenig and Melie-Garcia 2010).

###### CPT EEG Pre Processing

Continuous Performance Task (CPT) EEG data were down sampled (1000Hz) and bandpass filtered (fourth-order, zero-phase, Butterworth filter, 0.1-100 Hz) and bandstop filtered (48-52 Hz; to remove 50Hz line noise). A custom function appended the Presentation .log file to the EEG data, labelling participant’s response accuracy to ‘Go’ and ‘No Go’ conditions. An automatic artefact rejection algorithm was used prior to manual artefact rejection. Adaptive Mixture of Independent Component Analysis (AMICA) (Palmer et al. 2011) was used to separate components, which were visually inspected to manually select and remove eye movements and remaining muscle activity artefacts. Data was re-referenced to average, epoched from -100 to 900ms and individually averaged to ‘Go and ‘No Go’ conditions.

A two-way ANOVA was used to compare number of included epochs across group and condition and demonstrated a main effect of condition (*F (1,36) =* 8.061, *p* = 0.006), but no main effect of group or interaction between group and condition (both p > 0.05). Total mean number of epochs for the ‘Go’ condition (*M=* 95, *SD =* 5.24) were significantly higher than the ‘No Go’ condition (*M=* 81, *SD =* 8.35).

###### RAGU Analysis

For statistical analysis of ERP’s, the Randomisation Graphical User Interface (RAGU), an open-source MATLAB based toolbox was utilised (Koenig et al. 2011). RAGU is a multivariate approach that uses powerful, assumption free, randomisation statistics to analyse multi-channel event related potential (ERP) data (Koenig et al. 2011). RAGU allows for comparisons of overall neural response strength (with the global field power - GFP test). Global brain activity can be described by the global field power (GFP), which is mathematically defined as the root of the mean of the squared potential differences at all *K* electrodes (i.e. *V_i_(t)*) from the mean of instantaneous potentials across electrodes (i.e. *V_mean_(t)*) (Lehmann and Skrandies 1980). A measure of the strength of the electric field over the brain at each point in time, GFP is representative of the global brain response to an event. Local maxima of GFP curve represents instances of strongest field strength and highest topographic signal to noise ratio (Khanna et al. 2015). RAGU also allows for comparisons of the distribution of neural activity between group and condition, using topographic analysis of variance (TANOVA). The TANOVA is a non-parametric randomisation test based on global dissimilarities between electric fields. In contrast to electrode-wise comparisons, the TANOVA computes global dissimilarity of the whole electrical field topographies between conditions or groups and tests for the significance of these topographic differences at each time point (Ruggeri et al. 2019). The TANOVA was implemented on the amplitude-normalized maps (GFP = 1), such that the results obtained are independent of variations in the global field strength. The rationale behind this approach is that it enables significant differences between conditions to be attributed to partially different sources of the evoked potential, and not to different strengths of similar source distributions. After observing periods above the duration threshold, post-hoc t-maps were produced to enable further investigation of the topographic distribution of the observed differences.

To protect the results from false positives caused by multiple testing, additional testing checked whether the duration of continuous periods of significance observed in our data exceeded the duration of significant periods in > 95% of the randomised data. This ensured the duration of a significant time period exceeded chance. Five thousand permutations were conducted with an alpha of *p* < .05.

###### TMS-EEG Pre-Processing

TMS-EEG data were epoched around the TMS pulse (-1000ms to 1000ms) and baseline corrected to the pre-TMS pulse period (-500ms to -50ms). Data around the large signal from the TMS pulse (-5 to 15ms) were removed and linearly interpolated. Data were downsampled from 10,000Hz to 1,000Hz. The epoched data from two time points (Pre_iTBS and Post_iTBS) were concatenated and pre-processed concurrently to provide more datapoints for artifact rejection (as ICA performs better with more data (Korats et al. 2012), but note that the files were later split so only the pre iTBS data was analysed). An initial round of independent component analysis (Hyvarinen and Oja 2000) (FastICA) was then performed to remove components containing any large residual TMS-evoked EMG artefacts. A bandpass filter (1–100 Hz) was then applied and line noise was removed using a bandstop filter (48–52 Hz). Data was again visually inspected and any remaining noisy epochs removed. Finally, a second round of FastICA was performed to eliminate any remaining components representing blink, decay and noise-related artefacts. Both rounds of component rejection following FastICA utilised a semi-automated artefact detection algorithm, based on a previous research (Rogasch et al. 2014) and using TESA toolbox as a guide (Rogasch et al. 2017). Components representing the following artefacts were removed; eye blinks and saccades (mean absolute z score of the two electrodes larger than 2.5), persistent muscle activity (high frequency power that is 60% of the total power), decay artefacts and other noise-related artefacts (one or more electrode has an absolute z score of at least 4).

#### 1. **Results**

*Clinical Measures*

Table S2. Clinical Information for Control and mTBI

|  | Control | mTBI | *t* | *p* | *d* |
| --- | --- | --- | --- | --- | --- |
| N | 26 | 30 |  |  |  |
| HADS Anxiety (mean (sd)) | 1.67 (2.13) | 4.48 (3.38) | -3.44 | 0.002** | 0.99 |
| HADS Depression (mean (sd)) | 1.67 (2.50) | 4.33 (3.93) | -2.87 | 0.010** | 0.81 |
| HADS Total Score (mean (sd)) | 3.33 (4.37) | 9.57 (7.46) | -3.61 | 0.001** | 1.02 |
| RPQ 3 items (mean (sd)) | 0.28 (0.74) | 2.73 (2.42) | -5.26 | < 0.001** | 1.37 |
| RPQ 13 items (mean (sd)) | 2.36 (3.12) | 12.13 (10.41) | -4.89 | < 0.001** | 1.27 |
| MFI General Fatigue (mean (sd)) | 7.25 (2.05) | 11.75 (3.77) | -4.63 | < 0.001** | 1.48 |
| MFI Physical Fatigue (mean (sd)) | 6.08 (1.62) | 11.33 (1.99) | -8.46 | < 0.001** | 2.89 |
| MFI Reduced Activity (mean (sd)) | 6.75 (2.77) | 11.46 (3.87) | -4.19 | < 0.001** | 1.40 |
| MFI Reduced Motivation (mean (sd)) | 6.67 (2.46) | 9.25 (2.63) | -2.90 | 0.008** | 1.02 |
| MFI Mental Fatigue (mean (sd)) | 7.00 (3.69) | 12.12 (3.70) | -3.92 | < 0.001** | 1.39 |

*Note.* HADS; Hamilton Anxiety and Depression Scale, MFI; Multidimensional Fatigue Inventory, RPQ; Rivermead Post Concussion Questionnaire

#### *Cortical Activity Measures*

###### Digit Span EEG Task

The Topographic Consistency Test (TCT) showed topographical consistency of neural activity for both groups and both time points for most periods until 450ms after the presentation of a digit (See Fig S1).

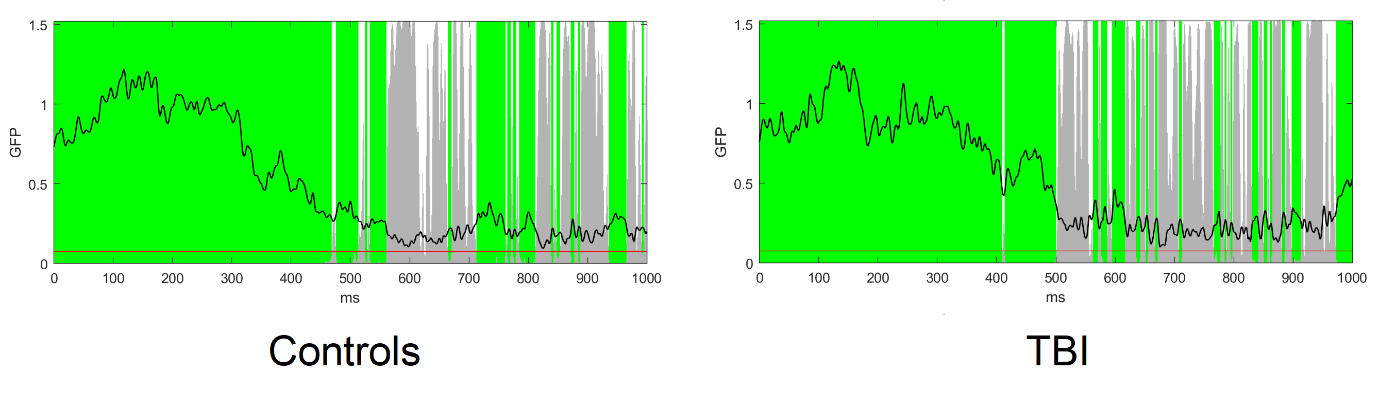

Fig S1. For the digit span task the TCT showed topographical consistency of neural activity for both groups until approximately 450ms after the presentation of a digit.

To examine potential differences in the distribution of neural activity between control and mTBI participants during the digit span EEG task Pre and Post iTBS, TANOVAs were conducted. Across the epoch (0-1000ms post digit presentation) no significant differences were demonstrated (all p > 0.05). Differences were detected in the GFP test in two significant time windows (617-655ms and 950-1000ms). However, these periods were during periods of topographical inconsistency in the TCT test, suggesting that any group differences during these periods may be the result of inconsistent neural activity patterns within groups, rather than a significant difference between the groups. As such, these differences were not interpreted further.

###### CPT EEG Task

The TCT showed topographical consistency of neural activity for both groups and both conditions for most time periods between 0 and 1000ms surrounding the presentation of a stimuli in the CPT task (See Fig S2).

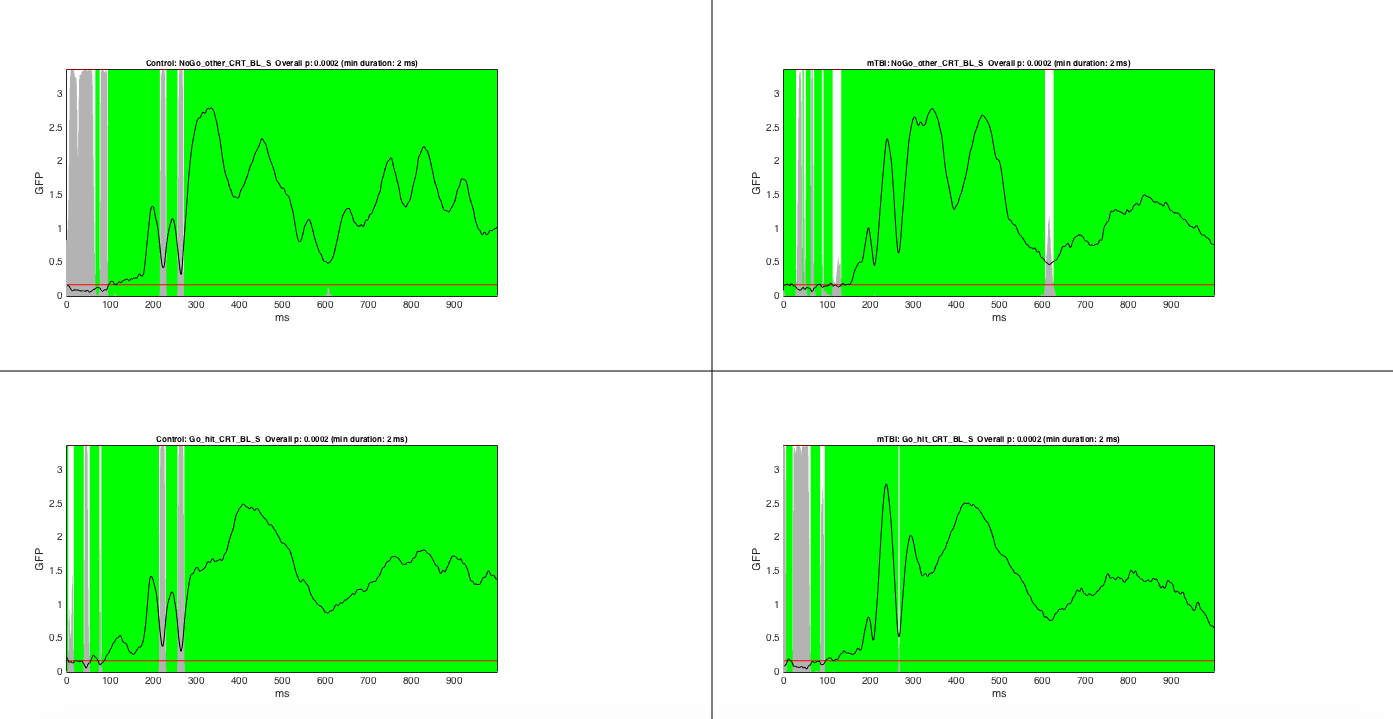

Fig S2. For the CPT task the TCT showed topographical consistency of neural activity for both groups and both conditions for most time periods between 0 and 1000ms following the presentation of a stimuli.

To examine potential differences in the distribution of neural activity between control and mTBI participants during the CPT task, TANOVA’s were conducted. A main effect of condition was demonstrated, a different distribution of neural activity to Go vs No Go conditions post stimulus presentation being shown (see Fig S3). Consistent with previous literature, these topographies demonstrate the typical Go and No Go neural responses, a NoGo-P300 potential maximal over fronto-central electrodes (Polich 2007), indicating that the task did generate the expected neural activity.

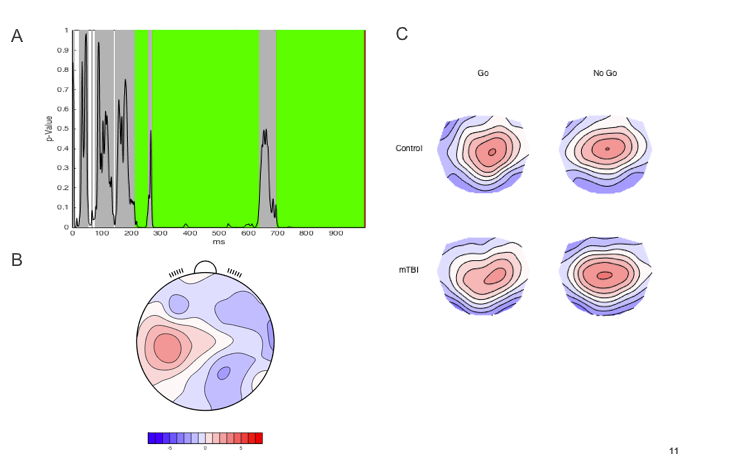

Fig S3.CPT neural results from RAGU. A—p values of the between-condition comparison for the real data against 5000 randomly shuffled permutations across the entire epoch (highlighted green sections reflect periods that exceed the duration control for multiple comparisons across time = 38 ms). B—t-map for NoGo topography minus Go topography. C—Averaged topographical maps for each group during the significant time window.

###### TMS-EEG

There was no significant difference between groups in resting motor threshold, *t* (55) = 1.51, *p* = 0.137.
